# Back to Basics: Correlates of HIV Risk in a Community Sample of Haiti

**DOI:** 10.1101/2020.10.10.20210476

**Authors:** Manisha Joshi, Guitele J. Rahill, Christopher Rice, Paul Phycien, Cameron Burris, Alyssa Maio

**Affiliations:** School of Social Work, College of Behavioral & Community Sciences, University of South Florida, Tampa, Florida, United States of America; School of Social Work, Florida International University. Miami, Florida, United States of America; OREZON Cité Soleil, Varreux, Port-au-Prince, Haiti, West Indies

**Keywords:** HIV knowledge, Personal HIV risk, Self-efficacy, Haiti

## Abstract

Haiti has a 2.2 % HIV prevalence (highest in the Caribbean); this has diminished from over 12% in the past three decades (depending on sex and gender, province, and neighborhood). Preliminary studies indicate that in the Cite Soleil neighborhood of Haiti (HIV prevalence >3%) as in socioeconomically equivalent adjacent neighborhoods, over 50% of girls and women experience non-partner sexual violence (NPSV), typically perpetrated by groups of men. Rates of NPSV against men in those neighborhoods were not available. Coercive sex heightens HIV risk. Accurate HIV knowledge empowers individuals (including survivors of NPSV) to assess personal HIV risk and increases likelihood of getting tested and of determining personal HIV status; thus, accurate HIV knowledge is foundational to behavioral risk reduction for victims in future consensual relationships and to engagement in either the HIV prevention or care continuum.

Between March and July 2017, we surveyed individuals 18 years or older (210 women, 257 men), assessing experience of NPSV, HIV knowledge, history of HIV testing, knowledge of HIV status, assessment of self-risk, and sexual risk behaviors. Nearly 30% of men and 24% of women endorsed having experienced NPSV. Knowledge of HIV transmission was low: 90% endorsed HIV myths, e.g. transmission occurs via public toilets, via sharing a glass with or by being exposed to a cough or sneeze from a person living with HIV. High endorsement of these myths contrasted with low endorsement of protective behavior: Only 14.3 % used a condom during consensual sex in the past year. Only 47.9% of the respondents had ever attended an HIV awareness program; 16% of knew their HIV status, although 79% assessed their HIV risk as moderate to high. Results regressing knowledge of HIV testing on participant characteristics indicated that women (OR=2.8), individuals with a partner (OR=2.2), individuals who attended an HIV awareness class (OR=2.1), individuals who knew someone with HIV (OR=3.9), and individuals who had an HIV test (OR=33.5) were more likely to know what an HIV test is. Participants who endorsed experiencing NPSV (OR=0.33) and those who had been diagnosed with an STI (OR=0.44) were less likely to know about HIV testing.

Experience of NPSV combined with low HIV knowledge, awareness and testing heighten the HIV prevention needs of Cite Soleil residents and underscore the need to return to basics on the road to HIV eradication in that context.

## Introduction

Current global narratives about the Human Immunodeficiency Virus (HIV) focus on the eradication of the virus by 2030 through engagement of persons living with HIV (PLHIV) in the care continuum (World Health Organization [WHO], 2018). In 2016, the United Nations set an agenda to end HIV and the Acquired Immune Deficiency Syndrome (AIDS) by 2020 (Joint United Nations Programme on HIV/AIDS [UNAIDS], 2017). To, successfully accomplish that goal, global targets include reducing the amount of new HIV infections to fewer than 500,000 globally and reducing the number of new infections in adolescent girls and young women to below 100,000 globally (UNAIDS, 2017-b). An additional goal is to guarantee that 90% of the people most at risk of becoming infected with HIV, including the key populations, can access HIV prevention measures (e.g. Pre-Exposure prophylaxis and Post-Exposure prophylaxes) (UNAIDS, 2017). These goals are not unrealistic, given that as of 2017, over 50% of adults and children living with HIV globally were engaged in treatment with antiretroviral drugs (ART) (WHO, 2018).

### Progress in the Battle to eradicate HIV in Haiti

Haiti has long been a prioritized nation for HIV prevention and treatment. With a population of approximately 11,000,000, HIV prevalence in is over 2.2%. Approximately 150,000 persons in Haiti were designated PLWHA, nearly 9,100 of whom were receiving ART, and 4,100 of whom died from AIDS-related complications (Centers for Disease Control and Prevention [CDC], 2018).

Over the past three decades, the CDC, the Joint United Nations Programme on HIV AIDS (UNAIDS) and Haiti-based health providers have successfully provided access to HIV testing and counseling to over a million people and engaged 60% of the nation’s estimated number of PLHIV in care and on ART (CDC, 2017). Accordingly, HIV prevalence in Haiti has decreased from 12% to 2.2% (Koenig et al., 2010; CDC, 2017). Nevertheless, the 2010 earthquake did reduce Haiti’s HIV prevention and care capacity, constraining HIV prevention efforts towards key target populations such as pregnant women, sex workers, prisoners, and men who have sex with men (MSM) (Ministère de la Santé Publique et de la population [MSPP]/The Haitian Ministry of Health, 2016). The inclusion of pregnant women as a key population for HIV prevention should ostensibly expose them to HIV knowledge, awareness, and testing, as means of preventing HIV in their consensual liaisons and in preventing transmission of the virus to their offspring. However, although, new HIV infections in the general population of Haiti and in the key populations reduced by 50% in 2016 (MSPP, 2016), data concerning HIV prevalence and knowledge of HIV status among key populations vary (UNAIDS, 2016).

### Feminization of HIV in Haiti

Available data on HIV prevalence in Haiti indicate that between 1.9 and 3.5% of Haiti’s 11 million people are PLHIV; these statistics fluctuate based on the region or neighborhood examined. What has remained consistent across reports is that girls and women in Haiti bear a heavier HIV burden than that borne by their male counterparts, a gender disparity referred to as the “feminization of HIV in the country” (MSPP, 2016, p. 26). Indeed, an early study that investigated HIV risk factors for Haitian adolescents and young adults in a disadvantaged urban location of Haiti reported that HIV was disproportionally present in its female participants and that HIV rates during adolescence and early adult years were greater for females (Dorjgochoo et al., 2009).

More recently, HIV prevalence rates among Haitian women aged 15-49 in 2017 were 2.7% greater than the national prevalence, in contrast to 1.7% among men (MSPP, 2017). MSPP recently acknowledged that the ratio of female to male HIV infection is 1:59 indicating that Haitian females are more vulnerable than Haitian males, regardless of the age range. The differential prevalence rate based on gender is grimmer within the 25-29 year old category, in which HIV females bear a burden of HIV that is four times greater than males. Moreover, the HIV rate among Haitian males has shown a reduction over the past several years while it has increased among females (MSPP, 2016).

In addition to gender disparities in HIV prevalence in Haiti, certain socioeconomically marginalized neighborhoods may not be counted in surveillance efforts, causing them to lag behind in engagement in the HIV prevention and care continua.

### Cité Soleil

Cité Soleil, population approximately 300,000, is a densely populated shantytown in Port-au-Prince. Its location on the ocean belies its extreme poverty. The average household in Cité Soleil earns less than $2.00 a day and comprises five or more persons living in cramped spaces (Marcelin, Page, & Singer, n. d.). There, hunger, unemployment, inadequate housing, and poor infrastructure such as lack of electricity are among the top problems that enable gang violence, gender-based violence and lack of legal remedies for victims. In addition, standing water and lack of consistent health providers at local clinics and hospitals result in a disproportionate burden of health conditions such as malaria and tuberculosis, such that most residents do not live past age 50, below Haiti’s average life expectancy of 52 (CDC, 2017; UNAIDS, 2014).

Roughly 52% of Cité Soleil residents are females, a significant proportion (over 50%) of which have experienced contextually brutal acts of non-partner sexual violence (NPSV), typically perpetrated by groups of men who aim to destroy their reproductive system (D’Adesky, & PotoFanm+Fi, 2012; Faedi, 2008; Joshi, Rahill, Lescano, & Jean, 2014). In a study of how NPSV and HIV risk intersect in Cité Soleil, none of the victims (self-named) (n=16) was aware that NPSV raised their HIV risk (Joshi et al., 2014). They also did not know how to prevent HIV; all of these women also reported that they would be safer if they had a male partner living in the home with them. Had they possessed accurate knowledge about how HIV is transmitted, they might have understood that the non-use of condoms by their multiple assailants did not simply pose a risk of pregnancy and social stigma associated with that pregnancy, but a risk to their sexual health and to the health of the children born of those acts of NPSV. Such knowledge might have led them to HIV testing and to early care. Rates of NPSV towards men is not available.

### HIV Knowledge

HIV knowledge, the ability to discern facts from myths about the transmission, diagnosis and treatment of HIV is an important part of HIV prevention (Swenson et al., 2010). Accurate HIV knowledge, i.e., being able to identify how HIV is transmitted and knowing about the importance of HIV testing, heightens HIV awareness, increases likelihood of getting tested, enables knowledge of personal HIV status, and is crucial to behavior change; all of these are foundational to risk reduction and engagement in either the HIV prevention or care continuum. Lack of knowledge about how HIV is transmitted results in underestimation of personal risk of acquiring or transmitting the virus. In fact, Haitian adolescents in the United States (US) who perceived themselves at higher risk for HIV were more likely to be HIV seropositive, and most were not aware of risk (Dorjgochoo, et al., 2009).

Notably, being able to discern HIV facts from myths does not automatically mean that one will engage in health promotion behaviors that mitigate risk, e.g., limiting one’s number of sexual partners; nor does it determine that one will correctly and consistently use condoms. Nonetheless, HIV knowledge is an important first step in behavior change in that it heightens awareness of risk, increases the likelihood of getting tested and thus of knowing one’s personal HIV status (Albarracín et al., 2005; Holtgrave & Curran, 2006).

The neighborhood violence in Cité Soleil hinders the entry of national surveillance experts and regional health providers, whose fear for their own safety and their attitudes towards Cité Soleil residents are obstacles to performing their assigned duties (Joshi, et al., 2014; Willman & Marcelin, 2010). Consequently, there is a dearth of reliable data concerning the HIV status of Cité Soleil residents, suggesting lack of HIV testing, which in turn requires HIV awareness, itself enhanced by HIV knowledge. This is important, because, colleagues at OREZON (Organisation pour la Renovation et l’Education du Zone Cité Soleil [Organization for the Renovation and Education of the Cité Soleil Zone) have long recognized the need for HIV prevention and treatment services for their constituents, who are victims of NPSV, since many succumb to AIDS before they realize they were seropositive for HIV (Placide, 2018).

To our knowledge, the last HIV study conducted in Cité Soleil was published nearly three decades ago, but although it did not evaluate HIV knowledge; it contributed that being female, between 20 to 29 years old, and having more than one sexual partner in the previous year, were correlated with HIV seropositivity (Boulos, et al., 1990).

### Purpose of the Study

The present work explores HIV knowledge, self-perception of HIV risk, behavioral risks, HIV awareness, and self-efficacy, and their correlates in a sample of Cité Soleil residents. We hope our study will clarify that there is a need in socioeconomically marginalized neighborhoods such as Haiti’s Cité Soleil, to return to HIV basics, i.e., implementing primary HIV education.

Understanding the correlates of this study participants’ HIV knowledge is important for several reasons. First, the study sample was drawn from a neighborhood that has an unknown burden of HIV/AIDS, that is situated in a country with the highest HIV prevalence in the Carribean region (UNAIDS, 2017a). Second, HIV knowledge can influence individuals to be tested, to gain knowledge of their HIV status. Knowledge of HIV status in turn, can contribute to individuals’ understanding of their HIV risk profile, thus, equipping them with information that informs their perceptions of self-efficacy, and ultimately to help them be safe in consensual relationships (Johnston Polacek, Hicks and Oswalt, 2007; DuRant, Ashworth, Newman, Gaillard, et al., 1992; Polacek, Hicks, & Oswalt, 2007). Third, individuals who do not know their HIV status can transmit new HIV infections; if they knew their status, they might engage in care and avoid the most detrimental consequences associated with late diagnoses and late care (Marks, Crepaz & Janssen, 2006). Finally, understanding the correlates HIV knowledge in this sample of Cité Soleil residents is a first step in informing what aspects of HIV knowledge are needed to ultimately influence HIV testing, enhance greater linkage, engagement, and retention in care and to increase rates of viral suppression in that community.

### Theoretical framework

We merged the Information-Motivation-Behavioral skills (IMB) model with gender vulnerability in this study. Although there is little evidence of its use in Haiti, there is strong evidence for the relevance of the IMB model in predicting health behaviors among Haitians in the United States (Malow, Stein, McMahan, Dévieux, Rosenberg & Jean-Gilles, 2008). In the IMB model, information refers to having knowledge about HIV transmission, that is, having behavior-related information and challenging misconceptions/myths regarding HIV that affect decision-making, and thus is foundational to behavior change (Chang, Choi, Kim, & Song, 2014). Motivation, in the IMB model, includes personal motivation, beliefs about testing or intervention outcomes, attitudes toward health behaviors, social motivation, and perceived social support for engaging in a specific behavior aimed at HIV risk reduction (Chang et al., 2014). Finally, behavioral skills in the IMB model are the skills necessary to perform a health behavior and emphasize both the enhancement of individuals’ objective skills and increased perceived self-efficacy when working to produce change (Chang et al., 2014). All three components of the IMB model are essential in eliciting behavior change.

Gendered vulnerability indicates that a girl or woman’s gender affects her ability to anticipate, deal with, and recover from sexual manipulation, sexual violence or unplanned pregnancies that might result from NPSV, due to socialization experiences that are formed and supported by traditional, cultural, socio-economic, and environmental factors (Enarson, 1998). Lamour (2018) draws from Enarson to posit differentiated roles of Haitian boys and girls, roles that are at the heart of all types of violence against girls and women in Haiti, especially in the slum regions. Specifically, she notes that from an early age, girls are socialized to remain physically and socially attached to the family and that Haitian parents instill in girls an ethic of responsibility for and accountability to others, as girls are considered a resource. Alternatively, Haitian boys are socialized as free selves who as adults will have the time and resources to engage in politics, and even their ideal of politics, governance and rule, does not model accountability to society. The application of IMB and gendered vulnerability in the present study means that Haitian female participants would possess less accurate HIV knowledge, less HIV awareness, would be less likely to engage in HIV prevention, would report less motivation or self-efficacy to engage in HIV prevention, would report lower rates of HIV testing, and would have less knowledge of their HIV status than their male counterparts.

## Methods

### Ethical Considerations

Our study, conducted between March and July 2017, was approved by our University Institutional Review Board and by the National Bioethics Committee of Haiti. Using a community-based participatory approach, we collaborated with OREZON. The OREZON staff are trained in human subjects’ protection, and contributed to the different aspects of this study, including the development and translation of the survey, recruitment of participants, data collection, and manuscript preparation.

### Inclusion Criteria

Inclusion criteria included 18 years of age or older, having no cognitive impairment that would preclude correct completion of the survey, and being willing and able to voluntarily participate in a study regarding the health of Cité Soleil residents. We attempted to recruit as many females as males.

### Recruitment and Sampling

Using purposive sampling procedures, including venue-based, snowball, and convenience sampling, we collected data in two phases. In Phase 1, we recruited from female attendees who participated in an International Women’s Day event in Cité Soleil. In Phase 2, male and female volunteers, who had heard about the survey via word-of-mouth or had seen this study’s fliers were recruited and consented by OREZON. The final sample size was 467 (n=257 males, 210 females).

### Measures

All study measures and consent forms were translated in Haitian *Kreyòl* by certified translators at OREZON Cité Soleil’s MEXSAF (Maison des Expressions sans Fontières [House of Languages without Borders]). One of the author who is a native Haitian and fluent in English as well as in Haitian *Kreyòl* back translated all study documents, insuring that they were consistent with the original documents.

Our survey consisted of questions about sociodemographic characteristics and basic questions that assessed HIV knowledge, distinguishing between accurate knowledge and misconceptions, history of HIV testing, knowledge of HIV status, assessment of self-risk and sexual risk behaviors. To avoid embarrassing participants who might be preliterate, we offered participants the opportunity of having the PA or an OREZON colleague read the questions aloud and write down the responses. A similar option was offered during the informed consent process. Participants’ ability to provide informed consent was confirmed by the PA and OREZON colleagues when participants correctly described the purpose of the study and the benefits and risks associated with the study.

We selected the most basic HIV knowledge questions our team could agree on, drawing from Heckmann and colleagues’ HIV Knowledge Scale and others (Carey, Morrison-Beady & Johnson, 1997; Denison, O’Reilly, Schmid, Kennedy, & Sweat, 2008; Gowen, Bovender & Edwards, 2016; Heckman, et al., 1995; Villegas, Cianelli, Gonzalez-Guarda, Kaelber, Ferrer, & Peragallo, 2013). For example, respondents answered either Yes or No, if they: had a current partner, had ever been diagnosed with a sexually transmitted infection (STI), had ever attended an HIV awareness class, and had ever been tested for HIV. They were also asked to distinguish the most effective way to prevent HIV from several options and to answer either Yes or No to if they: knew what an HIV test is, had ever been tested for HIV, knew their HIV status, knew persons with HIV, used condoms each time they had consensual sex in the previous 12 months. If they used condoms in the previous 12 months, they were asked whose idea it was, i.e., theirs or a partner’s. Additionally, we asked them to rate their personal risk of having HIV, and to answer Yes or No to statements that are widely considered HIV myths (i.e., if the virus can be transmitted via kissing someone on the cheek, by sharing a glass with someone who has HIV, by having someone sneeze or cough on them, and by using public toilets, the most effective way to prevent HIV).

### Analysis

We calculated descriptive statistics (i.e., frequencies, percentages) to analyze male and female participants’ responses to questions about socio-demographics, HIV knowledge and awareness, self-assessment of HIV risk, behavioral risks, and self-efficacy and preventions efforts. We then conducted bivariate analyses (chi-square tests) to explore associations between five specific HIV awareness variables: 1) knowing the most effective method of preventing HIV (abstinence or other); 2) knowing what is an HIV test (Yes or No); 3) whether a condom was used every time during consensual sex in last 12 months (Yes or No),; 4) if condom was used, whose idea was it to use it (My idea or Not my idea); and, 5) the participants’ personal HIV risk rating (very low risk or Moderate/Very high risk) and participant characteristics of – gender, age, education, employment, having a partner, having a child, having ever been a victim of NPSV, having ever attended an HIV awareness class, having ever been diagnosed with and STI, having ever had an HIV test, knowing one’s HIV status, and knowing persons’ with HIV. We also calculated the effect sizes for these analyses using Cramer’s *V*.

We then explored to what degree participant’s personal characteristics were linear functions of each of five outcomes indicating HIV awareness: 1) abstinence as the most effective HIV prevention method; 2) knowing what an HIV test is (as indicators of HIV knowledge and awareness); 3) using condom every time during sex in past 12 months; 4) taking the lead in negotiating use of condom (as indicators of self-efficacy and prevention efforts); and, 5) rating of moderate/high personal HIV risk (as an indicator of self-assessment of risk) For each outcome, we limit inclusion of personal characteristics to those found statistically significant in the bivariate analysis.

Because of the binary nature of each of these five outcome variables, we employed logistic regressions. Standard diagnostic statistics were calculated before undertaking logistic regressions (e.g., Variance Inflation Factor) and were found to be acceptable for each of the five regression models.

## Results

Socio-demographic characteristics of survey respondents are presented in Table 1. More than 80% of all respondents were 35 years or less, 55% were males, and about 40% of both males and females were in paid employment (X^2^ = 2.61; p=.27). A significantly higher percentage of females as compared to males had attended secondary school or higher (20% vs. 8.27%) while a higher percentage of males reported either no school or preschool (23.35% vs. 11.90%) (X^2^ =20.30; p<.001). A significantly higher percentage of females reported having a partner (67.14%) in comparison to males (30.74%) (X^2^ = 61.46; p<.001). Half of both males (55.25%) and females (58.10%) indicated that they had children (X^2^ = 0.37; p=.053). About a quarter of all respondents (26.98%) had experienced NPSV in their life, with males reporting a slightly higher percentage (29.57%) than females (23.81%) (X^2^ = 1.96; p=.16).

**TABLE 1.** Characteristics of Participants by Gender (N=467)

| Survey items | Total | Men %<br>(n=257) | Women %<br>(n=210) | $X^2$ | $df$ | $p$ |
| --- | --- | --- | --- | --- | --- | --- |
| Age |  |  |  | 2.61 | 2 | 0.27 |
| 18-25 | 41.11 | 42.02 | 40.00 |  |  |  |
| 26-35 | 41.97 | 43.58 | 40.00 |  |  |  |
| 36 and higher | 16.92 | 14.40 | 20.00 |  |  |  |
| Educational status |  |  |  | 20.30 | 2 | **0.00** |
| None/preschool | 18.20 | 23.35 | 11.90 |  |  |  |
| Primary to 8<sup>th</sup> grade equivalent | 68.31 | 68.48 | 68.10 |  |  |  |
| Secondary and higher | 13.49 | 8.17 | 20.00 |  |  |  |
| Paid for work/Employment |  |  |  | 0.48 | 1 | 0.48 |
| Yes | 41.76 | 43.19 | 40.00 |  |  |  |
| No | 58.24 | 56.81 | 60.00 |  |  |  |
| Have a boyfriend/girlfriend/partner |  |  |  | 61.46 | 1 | **0.00** |
| Yes | 47.11 | 30.74 | 67.14 |  |  |  |
| No | 52.89 | 69.26 | 32.86 |  |  |  |
| Have children |  |  |  | 0.37 | 1 | 0.53 |
| Yes | 56.53 | 55.25 | 58.10 |  |  |  |
| No | 43.47 | 44.75 | 41.90 |  |  |  |
| Ever been a victim of sexual violence |  |  |  |  |  |  |
| Yes | 26.98 | 29.57 | 23.81 | 1.94 | 1 | 0.16 |
| No | 73.02 | 70.43 | 76.91 |  |  |  |

Survey responses related to participants’ HIV knowledge and awareness, behavioral risks, self-assessment of HIV risk, and self-efficacy and prevention efforts are presented in Table 2. A substantially higher percentage of females than males had attended an HIV awareness class in their life (72.86% vs. 27.63%; X^2^ = 94.72; p<.001), knew what an HIV test is (76.67% vs. 35.80%; X^2^ = 77.15; p<.001), ever had been tested for HIV (70.95% vs. 35.41%; X^2^ = 58.44; p<.001), and knew people with HIV (24.76% vs. 14.79%; X^2^ = 7.39; p=.007). For questions that shed light on participants’ ability to discern HIV facts from myths [i.e., if the virus can be transmitted via kissing someone on the cheek (X^2^ = 0.849; p=.357), by sharing a glass with someone who has HIV (X^2^ = 1.31; p=.251), by having someone sneeze or cough on them (X^2^ = 13.09; p=.001), and by using public toilets (X^2^ = 8.51; p=.004)], about 90% or more of both males and females provided the wrong answers.

**TABLE 2.** HIV Awareness and Knowledge, Self-assessment of HIV Risk, Behavioral Risks, Self-efficacy, and Prevention Efforts, by Gender.

| | Total | Men %<br>(n=257) | Women %<br>(n=210) | $X^2$ | df | p |
| --- | --- | --- | --- | --- | --- | --- |
| **HIV awareness** |  |  |  |  |  |  |
| Ever attended HIV awareness program/class |  |  |  | 94.72 | 1 | **0.000** |
| Yes | 47.97 | 27.63 | 72.86 |  |  |  |
| No | 52.03 | 72.37 | 27.14 |  |  |  |
| Know what is an HIV test |  |  |  | 77.75 | 1 | **0.000** |
| Yes | 54.18 | 35.80 | 76.67 |  |  |  |
| No | 45.82 | 64.20 | 23.33 |  |  |  |
| Ever had an HIV test in your life |  |  |  | 58.44 | 1 | **0.000** |
| Yes | 51.39 | 35.41 | 70.95 |  |  |  |
| No | 48.61 | 64.59 | 29.05 |  |  |  |
| Know your HIV status |  |  |  | 1.47 | 1 | 0.224 |
| Yes | 16.27 | 14.40 | 18.57 |  |  |  |
| No | 83.73 | 85.60 | 81.43 |  |  |  |
| Know someone with HIV |  |  |  | 7.39 | 1 | **0.007** |
| Yes | 19.27 | 14.79 | 24.76 |  |  |  |
| No | 80.73 | 85.21 | 75.24 |  |  |  |
| **HIV knowledge: Ability to discern HIV facts from myths** |  |  |  |  |  |  |
| Can get HIV by kissing someone who has HIV on cheek |  |  |  | 0.849 | 1 | 0.357 |
| Yes | 95.72 | 96.50 | 94.76 |  |  |  |
| No | 4.28 | 3.50 | 5.24 |  |  |  |
| Can get HIV by sharing a drink glass with someone with HIV |  |  |  | 1.31 | 1 | 0.251 |
| Yes | 92.93 | 94.16 | 91.43 |  |  |  |
| No | 7.07 | 5.84 | 8.57 |  |  |  |
| Can get HIV from HIV+ person coughing/sneezing on you |  |  |  | 13.09 | 1 | **0.000** |
| Yes | 94.65 | 98.05 | 90.48 |  |  |  |
| No | 5.35 | 1.95 | 9.52 |  |  |  |
| Can get HIV by using public toilets |  |  |  | 8.51 | 1 | **0.004** |
| Yes | 92.51 | 95.72 | 88.57 |  |  |  |
| No | 7.49 | 4.28 | 11.43 |  |  |  |
| Most effective way of preventing HIV |  |  |  | 4.22 | 1 | **0.040** |
| Abstinence | 42.83 | 47.08 | 37.62 |  |  |  |
| Other ways (e.g., being in a committed relationship) | 57.17 | 52.92 | 62.38 |  |  |  |
| **Self-assessment of HIV risk** |  |  |  |  |  |  |
| Rate your personal risk of getting HIV |  |  |  | 8.72 | 1 | **0.003** |
| Very low | 20.56 | 15.56 | 26.67 |  |  |  |
| Moderate/Very high | 79.44 | 84.44 | 73.33 |  |  |  |
| **Behavioral risks** |  |  |  |  |  |  |
| Ever been diagnosed with an STI |  |  |  | 46.03 | 1 | **0.000** |
| Yes | 37.47 | 23.74 | 54.29 |  |  |  |
| No | 62.53 | 76.26 | 45.71 |  |  |  |
| **Self-efficacy and prevention efforts** |  |  |  |  |  |  |
| Used condom each time during consensual sex in past year |  |  |  | 27.80 | 1 | **0.000** |
| Yes | 14.35 | 6.61 | 23.81 |  |  |  |
| No | 85.65 | 93.39 | 76.19 |  |  |  |
| If used condoms during sex in last 12 months, whose idea was it |  |  |  | 4.88 | 1 | **0.027** |
| My idea | 28.69 | 24.51 | 33.81 |  |  |  |
| Not my idea | 71.31 | 75.49 | 66.19 |  |  |  |

About 47.08% of males and 37.62% of females correctly reported that abstinence is the most effective HIV prevention method (X^2^ = 4.22; p=.040). Over half of females (54.29%) reported that they had received an STI diagnosis in comparison to a quarter of males (23.74%) (X^2^ = 46.03; p<.001). With reference to self-assessment of HIV risk, females and males were different: 26.67% females rated their risk is very low in comparison to 15.56% of males (X^2^ = 8.72; p=.003). Differences were also apparent in self-efficacy and prevention efforts of females and males. About a quarter of females (23.81%) versus only 6.61% of males reported that a condom was used each time they had consensual sex in the last 12 months (X^2^ = 27.80; p<.001). A higher percentage of females also indicated that when a condom was used, it was they who initiated the idea (33.81% females vs. 25.51% males) (X^2^ = 4.88; p=.027).

### Bivariate Analyses

Our bivariate analyses identified several characteristics that differentiated those who indicated - abstinence as the most effective HIV prevention method, knew what an HIV test is, used condom each time during sex in the past year, said it was their idea to use a condom, condom use, and gave themselves a moderate/very high HIV risk rating – from those who did not (Table 3). We found a significant difference between those who indicated yes-abstinence versus other for gender (X^2^ = 4.22; p=.040), education (X^2^ = 10.49; p=.005), having children (X^2^ = 5.07; p=.024), being a victim of NPSV (X^2^ = 39.85; p<.001), and having ever received an STI diagnosis (X^2^ = 8.33; p=.004). Among females, 37.62% correctly identified abstinence as the most effective method as compared to 47.08% for males. More than two-thirds of respondents (69.41%) who reported no education or preschool, did not choose abstinence as the correct answer. Even among those who reported secondary or higher education, a substantial minority (42.86%) provided the wrong answer. Having ever experienced NPSV and having received a diagnosis of STI were both inversely associated with an affirmative response to abstinence; 80.95% of those who reported NPSV and 65.71% of those who reported that they had an STI diagnosis provided the wrong answer for this question.

**TABLE 3.** Bivariate Analysis of Survey Data from Male Residents (n=257) and Female Residents (n=210) of Cité Soleil Slum in Haiti.

|  | Most effective way to prevent HIV, % |  |  |  |  | Know what is an HIV test, % |  |  |  |  | Used condom each time during sex in past year, % |  |  |  |  | Whose idea was it to use condom, % |  |  |  |  | Rate your personal risk of getting HIV, % |  |  |  |  |
| --- | --- | --- | --- | --- | --- | --- | --- | --- | --- | --- | --- | --- | --- | --- | --- | --- | --- | --- | --- | --- | --- | --- | --- | --- | --- |
|  | Abstinence | Other | X<sup>2</sup> | P | V | Yes | No | X<sup>2</sup> | P | V | Yes | No | X<sup>2</sup> | P | V | My Idea | Not My Idea | X<sup>2</sup> | P | V | Very low | Moderate or High | X<sup>2</sup> | P | V |
| Gender |  |  | 4.22 | **0.040** | 0.09 |  |  | 77.75 | **0.000** | -0.41 |  |  | 27.81 | **0.000** | -0.24 |  |  | 4.88 | **0.027** | -0.10 |  |  | 8.72 | **0.003** | 0.14 |
| Male | 47.08 | 52.92 |  |  |  | 35.80 | 64.20 |  |  |  | 6.61 | 93.39 |  |  |  | 24.51 | 75.49 |  |  |  | 15.56 | 84.44 |  |  |  |
| Female | 37.62 | 62.38 |  |  |  | 76.67 | 23.33 |  |  |  | 23.81 | 76.19 |  |  |  | 33.81 | 66.19 |  |  |  | 26.67 | 73.33 |  |  |  |
| Age |  |  | 2.36 | 0.300 | 0.07 |  |  | 0.35 | 0.837 | 0.03 |  |  | 0.26 | 0.878 | 0.02 |  |  | 2.00 | 0.367 | 0.06 |  |  | 3.15 | 0.207 | 0.08 |
| 18-25 | 46.88 | 53.13 |  |  |  | 52.60 | 47.40 |  |  |  | 13.54 | 86.46 |  |  |  | 26.56 | 73.44 |  |  |  | 22.40 | 77.60 |  |  |  |
| 26-35 | 40.82 | 59.18 |  |  |  | 55.61 | 44.39 |  |  |  | 15.31 | 84.69 |  |  |  | 32.14 | 67.86 |  |  |  | 16.84 | 83.16 |  |  |  |
| 36 and higher | 37.97 | 62.03 |  |  |  | 54.43 | 45.57 |  |  |  | 13.92 | 86.08 |  |  |  | 25.32 | 74.68 |  |  |  | 25.32 | 74.68 |  |  |  |
| Educational status |  |  | 10.49 | **0.005** | 0.15 |  |  | 9.02 | **0.011** | 0.14 |  |  | 3.02 | 0.221 | 0.08 |  |  | 0.14 | 0.930 | 0.02 |  |  | 0.13 | 0.937 | 0.02 |
| None/Preschool | 30.59 | 69.41 |  |  |  | 41.18 | 58.82 |  |  |  | 10.59 | 89.41 |  |  |  | 27.06 | 72.94 |  |  |  | 20.00 | 80.00 |  |  |  |
| Primary-8<sup>th</sup> grade | 43.26 | 56.74 |  |  |  | 55.49 | 44.51 |  |  |  | 14.11 | 85.89 |  |  |  | 29.15 | 70.85 |  |  |  | 20.38 | 79.62 |  |  |  |
| Secondary & higher | 57.14 | 42.86 |  |  |  | 65.08 | 34.92 |  |  |  | 20.63 | 79.37 |  |  |  | 28.57 | 71.43 |  |  |  | 22.22 | 77.78 |  |  |  |
| Paid for work |  |  | 3.95 | 0.050 | 0.09 |  |  | 2.65 | 0.104 | -0.08 |  |  | 2.60 | 0.107 | 0.07 |  |  | 0.16 | 0.685 | -0.02 |  |  | 26.30 | **0.000** | 0.24 |
| Yes | 48.21 | 51.79 |  |  |  | 49.74 | 50.26 |  |  |  | 17.44 | 82.56 |  |  |  | 27.69 | 72.31 |  |  |  | 9.23 | 90.77 |  |  |  |
| No | 38.97 | 61.03 |  |  |  | 57.35 | 49.74 |  |  |  | 12.13 | 87.87 |  |  |  | 29.41 | 70.59 |  |  |  | 28.68 | 71.32 |  |  |  |
| Have a partner |  |  | 0.95 | 0.328 | -0.05 |  |  | 69.52 | **0.000** | 0.39 |  |  | 52.65 | **0.000** | 0.34 |  |  | 21.98 | **0.000** | 0.22 |  |  | 9.99 | **0.002** | -0.15 |
| Yes | 40.45 | 59.55 |  |  |  | 74.55 | 25.45 |  |  |  | 26.82 | 73.18 |  |  |  | 39.09 | 60.91 |  |  |  | 26.82 | 73.18 |  |  |  |
| No | 44.94 | 55.06 |  |  |  | 36.03 | 63.97 |  |  |  | 3.24 | 96.76 |  |  |  | 19.43 | 80.57 |  |  |  | 14.98 | 85.02 |  |  |  |
| Have children |  |  | 5.07 | **0.024** | 0.10 |  |  | 0.14 | 0.711 | 0.02 |  |  | 0.09 | 0.765 | 0.01 |  |  | 2.24 | 0.135 | 0.07 |  |  | 0.40 | 0.528 | -0.03 |
| Yes | 47.35 | 52.65 |  |  |  | 54.92 | 45.08 |  |  |  | 14.77 | 85.23 |  |  |  | 31.44 | 68.56 |  |  |  | 21.59 | 78.41 |  |  |  |
| No | 36.95 | 63.05 |  |  |  | 53.20 | 46.80 |  |  |  | 13.79 | 86.21 |  |  |  | 25.12 | 74.88 |  |  |  | 19.21 | 80.79 |  |  |  |
| Ever been NPSV victim |  |  | 39.85 | **0.000** | -0.29 |  |  | 14.60 | **0.000** | -0.18 |  |  | 0.76 | 0.385 | 0.04 |  |  | 2.72 | 0.099 | -0.08 |  |  | 2.48 | 0.116 | -0.07 |
| Yes | 19.05 | 80.95 |  |  |  | 39.68 | 60.32 |  |  |  | 16.67 | 83.33 |  |  |  | 23.02 | 76.98 |  |  |  | 25.40 | 74.60 |  |  |  |
| No | 51.61 | 48.39 |  |  |  | 59.53 | 40.47 |  |  |  | 13.49 | 86.51 |  |  |  | 30.79 | 69.21 |  |  |  | 18.77 | 81.23 |  |  |  |
| Attended Awareness Class |  |  | 3.55 | 0.059 | 0.09 |  |  | 114.84 | **0.000** | 0.50 |  |  | 9.83 | **0.002** | 0.15 |  |  | 5.77 | **0.016** | 0.11 |  |  | 11.75 | **0.001** | -0.16 |
| Yes | 47.32 | 52.68 |  |  |  | 79.91 | 20.09 |  |  |  | 19.64 | 80.36 |  |  |  | 33.93 | 66.07 |  |  |  | 27.23 | 72.77 |  |  |  |
| No | 38.68 | 61.32 |  |  |  | 30.45 | 69.55 |  |  |  | 9.47 | 90.53 |  |  |  | 23.87 | 76.13 |  |  |  | 14.40 | 85.60 |  |  |  |
| Know your HIV status |  |  | 1.33 | 0.249 | -0.05 |  |  | 8.85 | **0.003** | 0.14 |  |  | 8.38 | **0.004** | 0.13 |  |  | 13.37 | **0.000** | 0.17 |  |  | 1.10 | 0.295 | -0.05 |
| Yes | 36.84 | 63.16 |  |  |  | 69.74 | 30.26 |  |  |  | 25.00 | 75.00 |  |  |  | 46.05 | 53.95 |  |  |  | 25.00 | 75.00 |  |  |  |
| No | 43.99 | 56.01 |  |  |  | 51.15 | 48.85 |  |  |  | 12.28 | 87.72 |  |  |  | 25.32 | 74.68 |  |  |  | 19.69 | 80.31 |  |  |  |
| Know someone with HIV |  |  | 3.19 | 0.074 | -0.08 |  |  | 35.32 | **0.000** | 0.28 |  |  | 19.19 | **0.000** | 0.20 |  |  | 6.97 | **0.008** | 0.12 |  |  | 6.09 | **0.014** | -0.11 |
| Yes | 34.44 | 65.56 |  |  |  | 82.22 | 17.78 |  |  |  | 28.89 | 71.11 |  |  |  | 40.00 | 60.00 |  |  |  | 30.00 | 70.00 |  |  |  |
| No | 44.83 | 55.17 |  |  |  | 47.48 | 52.52 |  |  |  | 10.88 | 89.12 |  |  |  | 25.99 | 74.01 |  |  |  | 18.30 | 81.70 |  |  |  |
| Ever had an HIV test |  |  | 0.00 | 0.968 | 0.00 |  |  | 255.25 | **0.000** | 0.74 |  |  | 19.15 | **0.000** | 0.20 |  |  | 51.72 | **0.000** | 0.33 |  |  | 3.08 | 0.080 | -0.08 |
| Yes | 42.92 | 57.08 |  |  |  | 90.00 | 10.00 |  |  |  | 21.25 | 78.75 |  |  |  | 43.33 | 56.67 |  |  |  | 23.75 | 76.25 |  |  |  |
| No | 42.73 | 57.27 |  |  |  | 16.30 | 83.70 |  |  |  | 7.05 | 92.95 |  |  |  | 13.22 | 86.78 |  |  |  | 17.18 | 82.82 |  |  |  |
| Ever diagnosed with STI |  |  | 8.33 | **0.004** | -0.13 |  |  | 27.22 | **0.000** | 0.24 |  |  | 12.36 | **0.000** | 0.16 |  |  | 17.48 | **0.000** | 0.19 |  |  | 0.88 | 0.347 | 0.04 |
| Yes | 34.29 | 65.71 |  |  |  | 69.71 | 30.29 |  |  |  | 21.71 | 78.29 |  |  |  | 40.00 | 60.00 |  |  |  | 18.29 | 81.71 |  |  |  |
| No | 47.95 | 52.05 |  |  |  | 44.86 | 55.14 |  |  |  | 9.93 | 90.07 |  |  |  | 21.92 | 78.08 |  |  |  | 21.92 | 78.08 |  |  |  |

We found significant differences between those who knew what an HIV test is versus those who did not for gender (X^2^ = 77.75; p<.001), education (X^2^ = 9.02; p=.011), having a partner (X^2^ = 69.52; p<.001), having been a victim of NPSV (X^2^ = 14.60; p<.001), having ever attended an HIV awareness class (X^2^ = 114.84; p<.001), knowing your HIV status (X^2^ = 8.85; p=.003), knowing someone with HIV (X^2^ = 35.32; p<.001), having ever had an HIV test (X^2^ = 255.25; p<.001), and having ever received an STI diagnosis (X^2^ = 27.22; p<.001). As high as three-fourth of females (76.67%) knew what an HIV tests is in comparison to only about one-third of males (35.80%). A higher percentage of respondents with secondary or higher level of education knew what an HIV test is (65.08%) as compared to those with primary to 8^th^ grade education (55.49%) or no education/preschool level (41.18%). Ever having experienced NPSV was inversely associated with knowledge of an HIV test, with 60.32% victims indicating no knowledge of what an HIV test is. Having a partner was positively associated with knowing what an HIV test is: 74.55% of those who had a partner also knew about HIV tests. In addition, having ever attended an HIV awareness class, knowing one’s HIV status, knowing someone with HIV, having ever had an HIV test, and having received an STI diagnosis were all significantly positively associated with knowing what an HIV test is.

For condom use, significant differences were found between those who said yes versus no for gender (X^2^ = 27.81; p<.001), having a partner (X^2^ = 52.65; p<.001), having ever attended an HIV awareness class (X^2^ = 9.83; p=.002), knowing your HIV status (X^2^ = 8.38; p=.004), knowing someone with HIV (X^2^ = 19.19; p<.001), having ever had an HIV test (X^2^ = 19.15; p<.001), and having ever received an STI diagnosis (X^2^ = 12.36; p<.001). Among females, about a quarter (23.81%) agreed that a condom was used every time during consensual sex in the last 12 months while only 6.61% males agreed to the same. Only about, a quarter of respondents with steady partners (26.82%) reported using a condom every during consensual sex in the last 12 months. Having ever attended an HIV awareness class, knowing one’s HIV status, knowing someone with HIV, having ever had an HIV test, and having received an STI diagnosis were all negatively associated with using a condom each time during sex in the last 12 months. About 81% of those who said they had attended an HIV awareness class reported that they did not use a condom each time they had consensual sex in the past year. Similarly, 75% of those who knew their HIV status, 71.11% of those who knew someone with HIV, 78.75% of those who had ever had an HIV test, and 78.29% of those who had been diagnosed with STI did not use condom each time.

We also found significant differences between those who said it was their idea to use condom as compared to those who said it was someone else’s idea for gender (X^2^ = 4.88; p=.027), having a partner (X^2^ = 21.98; p<.001), having ever attended an HIV awareness class (X^2^ = 5.77; p=.016), knowing your HIV status (X^2^ = 13.37; p<.001), knowing someone with HIV (X^2^ = 6.97; p=.008), having ever had an HIV test (X^2^ = 51.72; p<.001), and having ever received an STI diagnosis (X^2^ = 17.48; p<.001). A higher percentage of females (33.81%) than males (24.51%) reported that they were the ones who initiated condom use. Of those who had a steady partner, 39.09% of respondents said it was their idea to use a condom as compared to 19.43% of respondents who said they do not have a partner. Other variables such as having ever attended an HIV awareness class, knowing one’s HIV status, knowing someone with HIV, having ever had an HIV test, and having received an STI diagnosis were all negatively associated with initiating condom use. For example, a majority of respondents (66.07%) who had reportedly attended an HIV awareness class in the past did not report that “condom use” was their idea. Even among those who knew someone with HIV, or had an STI diagnosis in the past, 60% said it was not their idea to use condom.

Finally, for personal HIV risk rating, significant differences emerged between those who rated very low risk as compared to moderate/high risk for gender (X^2^ = 8.72; p=.003), paid employment (X^2^ = 26.30; p<.001), having a partner (X^2^ = 9.99; p=.002), having ever attended an HIV awareness class (X^2^ = 11.75; p=.001), and knowing someone with HIV (X^2^ = 6.09; p=.014). Among females, a little over one-fourth (26.67%) rated their HIV risk level as very low as compared to 15.56% males. Being in paid employment, having a partner, having attended an HIV awareness class, and knowing persons with HIV were all positively associated with a moderate/high HIV risk rating: 90.77% of those who had paid employment, 73.18% of those with a partner, 72.77% of those who had attended an awareness class and 70% of those who knew someone with HIV rated themselves as at moderate/high risk of HIV.

For four variables that indicated participants’ ability to discern HIV facts from myths (i.e., if the virus can be transmitted via kissing someone on the cheek, by sharing a glass with someone who has HIV, by having someone sneeze or cough on them, and by using public toilets, the most effective way to prevent HIV), bivariate analyses were not conducted; the variation in each of these variables was insufficient to justify any meaningful analyses.

### Regression Analyses

The summary measures of model fit indicated that our five multiple logistic regression models were acceptable (see Table 4). For the first outcome variable (abstinence as most effective method versus others), females (vs. males; odds ratio [OR]=0.63; 95% confidence Interval [CI]=0.41, 0.96); those with no education or preschool education (vs. primary-eight grade; OR=0.57; 95% CI=0.33, 0.98), NPSV victims (vs. non-victims; OR=0.23; 95% CI=0.13, 0.38), and those with an STI diagnosis (vs. no STI diagnosis; OR=0.52; 95% CI=0.34, 0.81) were less likely to identify abstinence as the correct answer. Participants who had attended secondary school or higher were almost two times as likely to answer “abstinence” as compared to those who had attended primary to eighth grade (OR=1.87; 95% CI=1.03, 3.39).

**TABLE 4.** Multiple Logistic Regression Analyses of Survey Data from Male Residents (n=257) and Female Residents (n=210) of Cité Soleil Slum in Haiti.

| Predictors (Reference Category) | Outcome Variables |  |  |  |  |  |  |  |  |  |
| --- | --- | --- | --- | --- | --- | --- | --- | --- | --- | --- |
|  | Abstinence most effective for HIV prevention |  | Know what is an HIV test |  | Used condom each time during sex in past year |  | My idea to use condom |  | Moderate/Very high personal HIV risk rating |  |
|  | OR (95% CI) | P | OR (95% CI) | P | OR (95% CI) | P | OR (95% CI) | P | OR (95% CI) | P |
| Female (vs. Male) | 0.63 (0.41, 0.96) | **0.034** | 2.85 (1.46, 5.56) | **0.002** | 2.63 (1.37, 5.04) | **0.004** | 0.88 (0.54, 1.44) | 0.620 | 0.73 (0.43, 1.24) | 0.251 |
| Educational status (vs. Primary-8<sup>th</sup> grade) | - | - | - | - | - | - | - | - | - | - |
| None/Preschool | 0.57 (0.32, 0.98) | **0.046** | 0.95 (0.41, 2.18) | 0.920 | - | - | - | - | - | - |
| Secondary & higher | 1.87 (1.03, 3.39) | **0.039** | 1.04 (0.42, 2.62) | 0.917 | - | - | - | - | - | - |
| Paid for work (vs. No) | - | - | - | - | - | - | - | - | 4.30 (2.44, 7.58) | **0.000** |
| Have a partner (vs. No) | - | - | 2.19 (1.11, 4.32) | **0.022** | 7.20 (3.14, 16.50) | **0.000** | 1.57 (0.95, 2.60) | 0.077 | 0.64 (0.38, 1.08) | 0.100 |
| Have children (vs. No) | 1.34 (0.89, 2.01) | 0.148 | - | - | - | - | - | - | - | - |
| Ever been NPSV victim (vs. No) | 0.23 (0.13, 0.38) | **0.000** | 0.33 (0.16, 0.67) | **0.002** | - | - | - | - | - | - |
| Attended HIV Awareness Class (vs. No) | - | - | 2.09 (1.07, 4.08) | **0.029** | 0.71 (0.37, 1.38) | 0.320 | 0.62 (0.37, 1.06) | 0.085 | 0.59 (0.34, 1.02) | 0.063 |
| Know your HIV status (vs. No) | - | - | 0.67 (0.29, 1.55) | 0.355 | 1.34 (0.65, 2.72) | 0.418 | 1.80 (1.01, 3.19) | **0.044** | - | - |
| Know someone with HIV (vs. No) | - | - | 3.96 (1.6, 9.75) | **0.003** | 2.19 (1.13, 4.24) | **0.019** | 1.07 (0.61, 1.88) | 0.796 | 0.61 (0.34, 10.07) | 0.089 |
| Ever had an HIV test (vs. No) | - | - | 33.52 (17.48, 64.25) | **0.000** | 1.56 (0.78, 3.09) | 0.204 | 4.59 (2.67, 7.89) | **0.000** | - | - |
| Ever diagnosed with an STI | 0.52 (0.34, 0.81) | **0.004** | 0.44 (0.22, 0.91) | **0.027** | 0.98 (0.54, 1.80) | 0.968 | 1.50 (0.93, 2.43) | 0.095 | - | - |
| **Model fit** | LR X<sup>2</sup>=68.32;<br>df=6; p<.001 |  | LR X<sup>2</sup>=344.38;<br>df=10; p<.001 |  | LR X<sup>2</sup>=77.89;<br>df=7; p<.001 |  | LR X<sup>2</sup>=69.80;<br>df=7; p<.001 |  | LR X<sup>2</sup>=49.86;<br>df=5; p<.001 |  |

For the second outcome variable (know what is an HIV test versus not know), female participants were almost three times as likely as males (OR=2.85; 95% CI=1.46, 5.56), and people with a partner were two times as likely as those without a partner to know what is an HIV test (OR=2.19; 95% CI=1.11, 4.32). Participants who reported that they had ever attended an HIV awareness class were more than two times as likely as those who had not attended an awareness class to know about HIV tests (OR=2.09; 95% CI=1.07, 4.04). Those who knew someone with HIV were almost four times as likely as those who did not know anyone with HIV to know what an HIV test is (OR=3.96; 95% CI=1.6, 9.75). If a person had ever had an HIV test, they were more than 33 times as likely as those who had never had an HIV test to know what an HIV test is. We also found that participants who had experienced NPSV were less likely to know what is an HIV test as compared to non-victims (OR=0.33; 95% CI=0.16, 0.67). Similarly, those who reported that they had received an STI diagnosis in the past were less likely to know what is an HIV test as compared to those who had not received such a diagnosis (OR=0.44; 95% CI=0.22, 0.91).

We found significant differences between those who used a condom each time during sex in the past year and those who did not for gender, having a partner, and knowing someone with HIV. Females were more than two times as likely as males to indicate that a condom was used each time during sex in the last twelve months (OR=2.63; 95% CI=1.37, 5.04). Those who had a partner were more than seven times as likely as were those without a partner to use a condom each time during sex in the last 12 months (OR=7.20; 95% CI=3.14, 16.50). Participants who knew persons with HIV were more than two times as likely as were those who did not know anyone with HIV to use a condom each time during sex in the last 12 months (OR=2.19; 95% CI=1.13, 4.24).

For our fourth outcome variable (i.e. my idea to use condom versus somebody else’s idea”), only two predictor variables significantly related to answering “My idea to use condom”. Those who had ever had an HIV test in their life were more than four times as likely as were those who had never had an HIV test to report that it was their idea to use a condom OR=4.59; 95% CI=2.67, 7.89). Participants who knew their HIV status were almost two times as likely as were those who did not know their status to say that it was their idea to use a condom (OR=1.80; 95% CI=1.01, 3.19).

For our final model, we found that the only variable that related significantly to a moderate/high HIV risk rating was paid employment. Those who reported that they had a job in which they were paid were more than four times as likely as were those without a paid employment to endorse a personal rating of moderate/high HIV risk (OR=4.30; 95% CI=2.44, 7.58).

## Limitations

First, since a non-probability sampling approach was used, therefore, one needs to exercise caution in generalizing the findings to all residents of Cité Soleil or other comparable socioeconomically marginalized neighborhoods in Haiti. Second, we did have participants who either could not read or had challenges in reading the survey questions. However, study staff helped them by reading the questions aloud to them, along with the response options. Another limitation is that we did not assess the HIV stigma in our sample, and we know that in a similar context, HIV stigma can inhibit HIV testing and HIV disclosure once status is known (Kalichman & Simbayi, 2003). Future studies should include questions on stigma. Future studies should also plan to include key target populations such as sexual and gender minorities to build on the findings of the present study.

## Discussion

The majority (83.7%) of individuals who completed our survey did not know their HIV status.. Even if these individuals had known their HIV status, they might not have obtained access to the scarce medical resources that would affect their wellness, nor avoided the adverse consequences associated with late diagnoses and care. To increase the number of persons at risk for HIV who receive voluntary HIV counseling and testing services (VCT), barriers to the VCT sites in Haiti must be removed. Promoting communication among sexual partners is an important element to include in prevention programs (Kershaw et al., 2006). The development of positive behavioral changes can be promoted through interventions that incorporate dramatization and role-playing exercise that demonstrate new social roles (Rotheram-Borus et al., 2009). Research has shown that transformational narratives to encourage positive behavior changes are associated with reduced HIV risk behaviors (Rotheram-Borus et al., 2009). Peer-led prevention programs for adolescents have proved effective in the promotion of safe sexual behaviors as well (Holschneider & Alexander, 2003). The implementation of these suggestions could reduce the diagnoses of new HIV infections, increase the number of persons who correctly know their HIV status and their HIV risk profile.

Clearly, HIV knowledge, history of HIV testing and knowledge of one’s status are not sufficient to mitigate HIV transmission. However, HIV knowledge is a predictor of self-efficacy (Villegas et al., 2013), and self-efficacy, a “sense of personal power to exercise control over certain behaviors,” is a strong predictor of HIV risk reduction (Holschneider & Alexander, 2003, p.35).

In addition to the IMB model, it is crucial to consider external factors and their effect on behavior change. These include “environmental and structural barriers to behavior change, social and community support, and maintenance over time” (Rotheram-Borus et al., 2009). Some barriers specific to our sample that affect behavior change are the extreme poverty in Cité Soleil, NPSV, social marginalization, and the effects of the 2010 earthquake in reducing available HIV prevention resources (Joshi, et al., 2014).

An important impediment to policy implementation important goal in relation to policy is inequalities, violence, and discrimination against women, HIV-infected individuals, and key populations (UNAIDS, 2017-b).

Haitian women, especially those who live in Haiti’s most disadvantaged neighborhoods, are among the most vulnerable to HIV/AIDS.

Thus, broad, societal change is necessary to improve the conditions in Cité Soleil. Such efforts to reduce violence, including sexual violence, could help prevent the spread of HIV in the community. In order for community-level interventions to be successful in Cité Soleil, they must incorporate respect for clients, and support the potential to shift social norms (Rotheram-Borus et al., 2009). It is important for individuals to be respected and have sense of self-efficacy if they are to be motivated toward behavioral change.

In general, one’s sense of power to control their actions and behaviors, or efficacy, has been identified as a strong predictor of risky sexual behavior (Holschneider & Alexander, 2003). Couture and colleagues (2010) found that intention to use condoms was influenced by previous diagnosis with a STI, suggesting that negative past experiences can have an impact on future behaviors as well (Couture, Soto, Akom, Joseph, & Zunzunegui, 2010).

There are many complex individual, family, and community factors that interact and contribute to the violence rampant in Cité Soleil (Willman & Marcelin, 2010). For example, children who are exposed to aggression in their homes will seek refuge in their communities (Willman et al., 2010). Positive social support provided by communities can function as protective factors against risk factors experienced in homes (Willman et al., 2010). However, if communities are also filled with violence, they are not able to offer needed support to youth (Willman et al., 2010). This deleterious situation in Cité Soleil puts youth in a position of hopelessness with little chance of developing positive behaviors (Willman et al., 2010). This vicious cycle represents more broad societal problems but is part of the context in which any policy designed to raise HIV prevention awareness and reduce HIV infections must operate.

## Data Availability

Data are available upon request.

## Acknowledgements

We wish to acknowledge the administration and staff of OREZON Cité Soleil for their support. A special note of thanks to the men and women who participated in our survey.

## References

Boulos, R., Halsey, N. A., Holt, E., Ruff, A., Brutus, J. R., Quinn, T. C. … Boulos, C. (1990). HIV-1 in Haitian women 1982-1988. The Cité Soleil/JHU AIDS Project Team. Journal of Acquired Immune Deficiency Syndrome, 3(7), 721–728.

Carey, M. P., Morrison-Beedy, D., & Johnson, B. T. (1997). The HIV-Knowledge Questionnaire: Development and evaluation of a reliable, valid, and practical self-administered questionnaire. AIDS Behaviors,1(1), 60–74. DOI: http://10.13072/midss.233

Centers for Disease Control and Prevention [CDC] (2018). CDC Division of Global HIV & TB Country Profile: Haiti. Retrieved from https://www.cdc.gov/globalhivtb/where-wework/Haiti.pdf

Chang, S. J., Choi, S., Kim, S., & Song, M. (2014). Intervention strategies based on information-motivation-behavioral skills model for health behavior change: A systematic review. Asian Nursing Research, 8(3), 172–181. DOI: 10.1016/j.anr.2014.08.002

Couture, M., Soto, J. C., Akom, E., Joseph, G., & Zunzunegui, M. (2010). Determinants of intention to use condoms among clients of female sex workers in Haiti. AIDS Care, 22(2), 253–262. DOI: 10.1080/09540120903111478

D’Adesky, A.C. & PotoFanm+Fi (2012). Beyond shock: Charting the landscape of sexual violence in post-quake Haiti: Progress, challenges & emerging trends 2010-2012. Retrieved from http://potofi.files.wordpress.com/2012/12/beyond-shock-abridged-version-haiti-gbv-progress-report-nov-2012.pdf

Denison, J. A., O’Reilly, K. R., Schmid, G. P., Kennedy, C. E., & Sweat, M. D. (2007). HIV voluntary counseling and testing and behavioral risk reduction in developing countries: A meta-analysis, 1990–2005. AIDS and Behavior, 12(3), 363–373. DOI: http://10.1007/s10461-007-9349-x

Dorjgochoo, T., Noel, F., Deschamps, M. M., Theodore, H., Dupont, W., Wright, P., … Pape, J. W. (2009). Risk factors for HIV infection among Haitian adolescents and young adults seeking counseling and testing in Port-au-Prince. Journal of Acquired Immune Deficiency Syndromes (JAIDS), 52(4), 498–508. DOI: http://10.1097/QAI.0b013e3181ac12a8

Enarson, E. (1998). Violence against women in disasters: a study of domestic violence programs in the US and Canada. Violence against Women, 5(7): 742–768.

Faedi, B. (2008). The double weakness of girls: Discrimination and sexual violence in Haiti. Stanford Journal of International Law, 44(1), 147-203. Retrieved from http://bi.galegroup.com/essentials/article/GALE%7CA182200367/4e0763d46a9167024e18388e189707b5?u=tamp44898

Haitian Ministry of Health [MSPP]. (2016). Le Programme National de Lutte contre le Sida.

Heckman, T. G., Kelly, J. A., Sikkema, K., Cargill, V., Norman, A., Fuqua, W., … Hoffmann, R. (1995). HIV risk characteristics of young adult, adult, and older adult women who live in inner-city housing developments: Implications for prevention. Journal of Womens Health,4(4), 397–406. DOI: http://10.1089/jwh.1995.4.397

Holschneider, S. O., & Alexander, C. S. (2003). Social and psychological influences on HIV preventive behaviors of youth in Haiti. Journal of Adolescent Health, 33(1), 31–40. DOI: http://dx.doi.org/10.1016/S1054-139X(02)00418-4

Joint United Nations Programme on HIV/AIDS [UNAIDS] (2018). 2017 Global HIV statistics [Fact sheet]. Retrieved from http://www.unaids.org/sites/default/files/media_asset/UNAIDS_FactSheet_en.pdf

Joint United Nations Programme on HIV/AIDS (UNAIDS) (2016, November 22). HIV prevention among key populations. Retrieved from http://www.unaids.org/en/resources/presscentre/featurestories/2016/november/20161121_keypops

Joint United Nations Programme on HIV/AIDS (UNAIDS) (2016). Factsheet 2016. Retrieved http://www.unaids.org/sites/default/files/media_asset/20150901_FactSheet_2015_en.pdf

Joint United Nations Programme on HIV/AIDS [UNAIDS] (2017). HIV prevention 2020 road map: Accelerating HIV prevention to reduce new infections by 75%. Retrieved from http://www.unaids.org/sites/default/files/media_asset/hiv-prevention-2020-road-map_en.pdf

Joint United Nations Programme on HIV/AIDS (UNAIDS) & Haitian Ministry of Public Health and Population (MSPP) (2016). Global AIDS response progress report [data file]. Retrieved from http://www.unaids.org/en/regionscountries/countries/haiti

Joshi, M., Rahill, G. J., Lescano, C., & Jean, F. (2014). Language of sexual violence in Haiti: Perceptions of victims, community-level workers, and health care providers. Journal of Health Care for the Poor & Underserved 25(4), 1623–1640. DOI: http://10.1353/hpu.2014.0172

Kalichman, S. C. & Simbayi, L. (2003). HIV testing attitudes, AIDS stigma, and voluntary counseling and testing in a Black township in Cape Town, South Africa. Sexually Transmitted Infections, 79, 442–447. DOI: 10.1136/sti.79.6.442

Kershaw, T. S., Small, M., Joseph, G., Theodore, M., Bateau, R., & Frederic, R. (2006). The influence of power on HIV risk among pregnant women in rural Haiti. AIDS and Behavior, 10(3), 309–318. DOI: http://10.1007/s10461-006-9072-z

Koenig, S., Ivers, L. C., Pace, S., Destine, R., Leandre, F., Grandpierre, R., … Pape, J.W. (2010). Successes and challenges of HIV treatment programs in Haiti: Aftermath of the earthquake. HIV Therapy, 4(2), 145–160. DOI: http://10.2217/hiv.10.6

Malow, R. M., Judity, S., Mcmahon, R. C., Devieux, J., Rosenberg, R., & Jean-Gilles, M. (2008). Effects of a culturally adapted HIV prevention intervention in Haitian youths. Journal of the Association of Nurses in AIDS Care, 20(2), 110–121. DOI: http://10.1037/e517802008-001

Lamour, S. (2008). Partir pour mieux s’enraciner ou retour sur la fabrique du poto-mitan en Haiti. Déjouer le silence, contre discours sur les femmes haïtiennes (Leave to better take root or areturn to the poto-mitan concept in Haiti: To break the silence, anti-women narrative in Haiti. Montréal, Remue-Ménage: PresUniq.

Marcelin, L., Page, J. B., & Singer, M. (n. d.). Cité Soleil Project. Interuniversity Institute for Research and Development (INURED): Official presentation of results. Grant number: S□HA700□07□GR□014. http://www.inured.org/uploads/2/5/2/6/25266591/cite_soleil_project.pdf

Rotheram-Borus M. J., Swendeman, D., Flannery, D., Rice, E., Adamson, D. M., & Ingram, B. (2009). Common factors in effective HIV prevention programs. AIDS and Behavior, 13(3), 399–408. DOI: http://10.1007/s10461-008-9464-3

Swenson, R. R., Rizzo, C. J., Brown, L. K., Vanable, P. A., Carey, M. P., Valois, R. F., … Romer, D. (2010). HIV knowledge and its contribution to sexual health behaviors of low-income African American adolescents. Journal of the National Medical Association, 102(12), 1173–1182.

Villegas, N., Cianelli, R., Gonzalez-Guarda, R., Kaelber, L., Ferrer, L., & Peragallo, N. (2013). Predictors of self-efficacy for HIV prevention among hispanic women in south Florida. Journal of the Association of Nurses in AIDS Care,24(1), 27–37. DOI: http://10.1016/j.jana.2012.03.004

Willman, A. & Marcelin, L. H. (2010). “If they could make us disappear, they would!” youth and violence in Cité Soleil, Haiti. Journal of Community Psychology, 38(4), 515–531. DOI: http://10.1002/jcop.20379a

